# Context-Aware Digital Phenotyping of Youth Mental Health Using Mobile Ecological Prospective Assessments of Smartphone Use

**DOI:** 10.1101/2025.08.24.25334320

**Authors:** Jamin Patel, Sheriff Tolulope Ibrahim, Tarun Reddy Katapally

**Affiliations:** DEPtH Lab, School of Health Studies, Faculty of Health Sciences, Western University, London, Ontario, Canada N6A 5B9; Department of Epidemiology and Biostatistics, Schulich School of Medicine and Dentistry, Western University, London, Ontario, Canada N6A 3K7; Children’s Health Research Institute, Lawson Health Research Institute, 750 Base Line Road East, Suite 300, London, Ontario, Canada N6C 2R5

**Keywords:** anxiety, citizen science, depression, digital phenotyping, ecological momentary assessment, precision prevention, smartphone use, suicidal ideation, youth mental health

## Abstract

**Background:** Youth mental disorders affect 12-14% of adolescents globally and remain underdiagnosed and undertreated. Digital phenotyping offers a scalable approach to real-time behavioural monitoring via smartphones, yet most studies rely solely on passive measures such as screen time, overlooking contextual factors.

**Methods:** This cross-sectional study was part of the Smart Platform, a digital citizen science initiative that engaged youth aged 13-21 years. Participants completed a baseline survey on sociodemographic characteristics and mental health (depression, anxiety, and suicidal ideation). Over the next seven days, context-aware digital phenotyping was conducted, defined as the collection of ecologically valid, time-stamped behavioural data from personal devices. This was implemented through mobile ecological prospective assessments (mEPAs) to capture self-reported smartphone use context, including activity type, location, and social setting. Multivariable logistic regression assessed associations between smartphone use context and mental health, adjusting for sociodemographic covariates.

**Results:** Eighty-four youth completed the baseline survey and at least one mEPA. A higher proportion of smartphone use at home was associated with lower odds of depression (OR=0.105, 95% CI: 0.028-0.276) and anxiety (OR=0.150, 0.053-0.345). A greater proportion of smartphone use while alone was associated with higher odds of depression (OR=3.802, 1.622-11.241), as was a greater proportion of time spent internet surfing (OR=2.663, 1.238-6.843). Longer duration of smartphone use outside the home was associated with higher odds of depression (OR=4.289, 1.443-16.579).

**Conclusion:** Context-aware smartphone metrics may offer more informative digital phenotyping indicators of youth mental health than duration alone, supporting integration of multi-context measures into early detection and precision prevention frameworks.

## 1. Introduction

Youth mental health is a significant public health concern, with global prevalence estimates of mental disorders ranging from 12.4% among 10-14 year olds to 14% among 15-19 year olds [1]. These disorders rank among the top ten causes of disease burden in adolescents, with anxiety and depression comprising a substantial share [2]. Despite this high burden, approximately 54% of adolescents with mental health needs do not receive treatment, often due to stigma, poor access to care, and limited service availability [3]. The global economic burden is significant, with estimated losses approaching USD 5 trillion, reflecting healthcare expenditures, reduced productivity, and the long-term consequences of untreated mental illness [4].

In response, the Lancet Commission on Global Mental Health and Sustainable Development has emphasized the need for prevention, early identification, and the integration of mental health into both routine services and digital systems [5]. Traditional monitoring approaches, such as clinical assessments and retrospective surveys, are typically infrequent and vulnerable to recall bias, potentially missing critical fluctuations in mental health between encounters [6–11]. Recent studies using digital citizen science approaches have shown that youth often overestimate their smartphone use when asked to recall it, compared to what is captured through real-time data from their devices [10,11]. Similarly, differences have been found in how youth report physical activity, with only real-time methods capturing meaningful patterns related to their social and daily lives [8,9].

Digital phenotyping through smartphones has emerged as a scalable approach for capturing real-time indicators of mental health in everyday environments [12]. It involves using data from personal devices, such as smartphones and smart wearables, to continuously monitor behavioural and contextual signals, combining both user-initiated inputs (active data) and information recorded automatically (passive data) [13]. Active data may include brief mood check-ins or self-reports, while passive data, such as geolocation, screen time, movement, and app usage, is collected without user effort [14]. These streams can be sampled at high frequency, sometimes every few seconds, enabling detection of short-term fluctuations in behaviour that might otherwise go unnoticed [15].

Smartphone use has emerged as a growing focus in digital phenotyping research due to its near-universal adoption, with over 80% of adolescents reporting owning or having regular access to a smartphone in high-income countries [16]. Various measures of smartphone use, such as total screen time, frequency of app use, or the number of device pickups, have been associated with symptoms of depression and anxiety in youth [17]. However, prior studies have primarily relied on passive smartphone data, which may reflect a wide range of underlying behaviours or social contexts that cannot be inferred solely from usage patterns [18–21]. For instance, high smartphone use may involve academic work or meaningful social connection, but it may also reflect passive scrolling or compulsive checking – each with potentially distinct implications for mental health [22,23].

Mobile ecological prospective assessments (mEPAs) offer a promising solution to this limitation by capturing self-reported contextual information, such as social setting, activity type, or space of engagement, in close temporal proximity to passive data collection from individuals’ devices [8,9]. Moreover, because mEPAs can be scheduled or triggered shortly after a behaviour is detected, they are more closely aligned with the experience than traditional retrospective surveys, which often require recall over extended periods [8–10]. For instance, Al-akshar et al. (2024) deployed daily evening mEPA prompts asking youth whether they had used their smartphones earlier that same day for specific activities such as internet surfing, texting, or gaming, and to report the duration of each activity and who they were with during that time [10].

While previous studies have used mEPAs to assess mood or mental health symptoms, none have applied them to examine the contextual characteristics of smartphone use in relation to youth mental health [24–26]. Thus, this study aimed to examine how contextual features of smartphone use, specifically social setting, activity type, and location of engagement, are associated with anxiety, depression, and suicidal ideation in youth.

## 2. Methods

### 2.1. Study Design

This study was conducted as part of the Smart Platform, a digital citizen science initiative supporting ethical population health surveillance, integrated knowledge translation, policy development, and real-time interventions [27–29]. Youth aged 13 to 21 from Regina, Saskatchewan, Canada, participated in a seven-day study period. Data collection included a cross-sectional baseline survey capturing self-reported mental health and sociodemographic characteristics, alongside context-aware digital phenotyping. Digital phenotyping refers to the moment-by-moment quantification of human behaviours in naturalistic settings using data generated from personal digital devices such as smartphones and smartwatches [13,18]. In this study, it was implemented in an active form through mEPAs, where participants self-reported details of their smartphone use context – including activity type, location, and social setting – directly within a custom-built mobile application. This approach differs from passive metrics such as total screen time by incorporating situational and social information, allowing for the capture of behavioural patterns with greater ecological validity and relevance to mental health research. The study received ethics approval through a harmonized review process from both the University of Regina and the University of Saskatchewan (REB #2017-29).

### 2.2. Recruitment

Twelve high schools in Regina were approached for recruitment, of which five agreed to participate. Eligibility criteria required that participants have no medical condition preventing physical activity and, for those under age 16, parental implied consent initiated one week prior to recruitment. The research team partnered with schools to deliver presentations to students in grades 8 through 12, introducing the study, demonstrating app features, and assisting with the application installation process. A total of 818 participants provided informed consent within the app and completed the baseline survey on the first day, which included self-reported mental health and sociodemographic items. Participants were eligible for inclusion in the present study if they completed the baseline mental health section and provided at least one day of smartphone use data via mEPAs.

### 2.3. Measures

#### 2.3.1. Mental Health (Outcome Variable)

Mental health symptoms were assessed using self-reported baseline survey items capturing depressive symptoms, generalized anxiety, and suicidal ideation. Depressive symptoms were assessed with the question: “During the past 12 months, did you ever feel so sad or hopeless almost every day for two weeks or more in a row that you stopped doing some usual activities?” Participants answered “Yes” or “No,” with affirmative responses indicating the presence of depressive symptoms. Suicidal ideation was measured with the item: “During the past 12 months, did you ever seriously consider attempting suicide?” Responses were dichotomized, with a “Yes” indicating the presence of suicidal ideation. Generalized anxiety symptoms were assessed using the Generalized Anxiety Disorder-2 instrument, which included two items assessing the frequency of (1) feeling nervous, anxious, or on edge, and (2) being unable to stop or control worrying, over the past two weeks. Responses ranged from 0 (“Not at all”) to 3 (“Nearly every day”), yielding a total score from 0 to 6. A score of ≥3 was used to indicate anxiety symptoms.

#### 2.3.2. Smartphone Use (Primary Independent Variable)

Smartphone use was measured daily from days one through eight via time-triggered mEPAs delivered through the custom-built smartphone application. Each prompt was delivered between 8:00 PM and 11:30 PM and expired at midnight. Participants were asked to identify which types of smartphone activities they engaged in by selecting from a list of check-all-that-apply options, including internet surfing (e.g., browsing Facebook, Instagram, YouTube, Reddit, or reading the news), texting, and gaming. For each selected activity, participants were asked to report the number of minutes spent, the location where the activity occurred (e.g., home, school, park, street, or community centre), and with whom the activity was done (e.g., alone, with friends, siblings, parents, or other social groups).

A proportional indicator of internet-based smartphone use was calculated by dividing the minutes spent on smartphone internet surfing by the total minutes spent across all smartphone activities (i.e., internet, gaming, and texting). The proportion of smartphone use at home was derived by dividing total minutes of smartphone use occurring at home by the sum of minutes reported across home and outside-home settings. For the social context, the proportion of smartphone use while alone was calculated by dividing minutes spent using a smartphone alone by the total of minutes spent either alone or with others.

#### 2.3.3. Sociodemographic Characteristics

Participants self-reported demographic information through the baseline survey. Age was reported as a continuous variable. Ethnicity was reported using a multi-select item with a wide range of options (e.g., First Nations, African, Canadian, European). For analysis, ethnicity was dichotomized into “Canadian/European” and “ethnic minority, mixed, or other.” Parental education was assessed as the highest level of education attained by any parent or guardian, including options ranging from elementary school to completed post-secondary, as well as “don’t know” and “does not apply.” This variable was dichotomized into “completed post-secondary education” (college or university diploma/degree) versus “incomplete or unknown.” Part-time job status was included as a binary variable based on a yes/no response.

### 2.4. Data and Risk Management

All data were securely encrypted and transmitted to a protected cloud server, and no personally identifiable information was collected. Youth retained full agency over their data and could pause or discontinue data collection at any point during the study. To further support privacy, app permissions were intentionally minimized, avoiding access to sensitive information such as contact lists. Unique anonymized identifiers were assigned to each device, and data transmission was restricted to occur only under secure conditions, such as when the device was connected to Wi-Fi. Additionally, participants could opt to upload data only while their phones were charging, minimizing battery drain and unintended data use. All identifiers were hashed using lightweight algorithms to ensure that individual devices could not be traced back to users.

### 2.5. Statistical Analysis

Descriptive statistics were reported using medians and interquartile ranges for continuous variables, with Wilcoxon rank-sum tests used to compare baseline and complete samples. Categorical variables were summarized as proportions and compared using chi-square tests. Two sets of multivariable logistic regression models were conducted separately for anxiety, depression, and suicidal ideation, adjusting for age, parental education, ethnicity, and employment status. The first set of models examined associations between smartphone use context and mental health symptoms, while the second focused on duration of use. Influential observations, identified using Cook’s distance, were removed. Although school-level clustering was initially modelled with a random intercept, minimal between-school variance supported a fixed-effects approach. P-values were adjusted using the Benjamini-Hochberg procedure to correct for multiple comparisons. All analyses were conducted in R version 4.4.1, with statistical significance set at p < 0.05.

## 3. Results

Of the 818 youth who enrolled in the study, 363 completed the baseline survey, and 84 participants provided complete data for both the baseline survey and at least one mEPA prompt. As shown in **Table 1**, the median age was lower in the final sample (15 years, IQR = 3.0) compared to the baseline complete sample (16 years, IQR = 2.5), and this difference was statistically significant (W = 12,841, p = 0.019). Regarding parental education, 50% of participants in both samples reported that their parent(s) had completed a university or college degree, with the other 50% indicating incomplete education (χ^2^ = 0.007, p = 0.931). Ethnicity distribution remained consistent between the groups, with 60% identifying as visible minority, mixed, or other, and 40% as Canadian or European (χ^2^ = 0.021, p = 0.884). Of the baseline sample, 40% reported having a part-time job, compared to 30% in the final sample; however, this difference was not statistically significant (χ^2^ = 0.878, p = 0.349). Anxiety symptoms were reported by 30% of the original sample and 40% of the complete sample (χ^2^ = 2.103, p = 0.156). Depressive symptoms were reported by 40% of participants in both samples (χ^2^ = 0.241, p = 0.623). Suicidal ideation was reported by 20% of the baseline sample and 30% of the final sample (χ^2^ = 1.470, p = 0.225).

**Table 1.** Baseline demographics summary and comparison of original and complete samples.

|  | Baseline complete<br>(N=363) | Baseline + mEPA<br>complete (N= 84) |  |  |
| --- | --- | --- | --- | --- |
| Continuous variables | Median (IQR) | Median (IQR) | W | p-value |
| Age (years) | 16 (2.5) | 15 (3) | 12841 | 0.019* |
| Categorical variables | N (Proportion) | N (Proportion) | $\chi^2$ (df) | p-value |
| Parental Education |  |  |  |  |
| Completed University/College | 179 (0.5) | 41 (0.5) | 0.007 | 0.931 |
| Incomplete | 184 (0.5) | 43 (0.5) |  |  |
| Ethnicity |  |  |  |  |
| Visible Minority/<br>Mixed/Other | 200 (0.6) | 47 (0.6) | 0.021 | 0.884 |
| Canadian/European | 163 (0.4) | 37 (0.4) |  |  |
| Part-time Job |  |  |  |  |
| Yes | 132 (0.4) | 26 (0.3) | 0.878 | 0.349 |
| No | 231 (0.6) | 58 (0.7) |  |  |
| Anxiety |  |  |  |  |
| No | 250 (0.7) | 51 (0.6) | 2.103 | 0.156 |
| Yes | 113 (0.3) | 33 (0.4) |  |  |
| Depression |  |  |  |  |
| No | 214 (0.6) | 47 (0.6) | 0.241 | 0.623 |
| Yes | 149 (0.4) | 37 (0.4) |  |  |
| Suicidal Ideation |  |  |  |  |
| No | 282 (0.8) | 62 (0.7) | 1.470 | 0.225 |
| Yes | 81 (0.2) | 22 (0.3) |  |  |

Participants reported varied patterns of smartphone use across different contexts (**Table 2)**. The median time spent on smartphones at home was 30.0 minutes per day (IQR = 59.5), while the median time spent using smartphones outside the home was 27.4 minutes (IQR = 83.1). The median proportion of total smartphone use spent at home was 0.5 (IQR = 1.0). For activity type, participants reported a median of 60.0 minutes per day spent on internet browsing (IQR = 89.8) and 20.0 minutes on gaming or texting (IQR = 50.0). Internet browsing comprised a median of 80.0% of total smartphone use (IQR = 0.4). With regard to social context, participants reported a median of 51.3 minutes (IQR = 105.0) of smartphone use while alone, compared to 14.0 minutes (IQR = 51.8) while with others. The median proportion of time spent using smartphones alone was 0.80 (IQR = 0.6).

**Table 2.** Descriptive statistics showing patterns of smartphone use across different contexts.

|  | **Median** | **IQR** | **Minimum** | **Maximum** |
| --- | --- | --- | --- | --- |
| **Location** |  |  |  |  |
| Home | 30.0 | 59.5 | 0 | 572.1 |
| Outside | 27.4 | 83.1 | 0 | 858.5 |
| Proportion | 0.5 | 1.0 | 0 | 1 |
| **Activity Type** |  |  |  |  |
| Internet Browsing | 60.0 | 89.8 | 0 | 858.5 |
| Gaming or Texting | 20.0 | 50.0 | 0 | 540.0 |
| Proportion | 0.8 | 0.4 | 0 | 1 |
| **Social Context** |  |  |  |  |
| Alone | 51.3 | 105 | 0 | 515.7 |
| With others | 14 | 51.8 | 0 | 960.0 |
| Proportion | 0.8 | 0.6 | 0 | 1 |

**Table 3** presents the results of logistic regression models examining associations between smartphone use context and mental health symptoms. After adjusting for age, ethnicity, parental education, part-time employment, and school, a higher proportion of smartphone use at home (vs. outside) was consistently associated with lower odds of both depressive symptoms (OR = 0.105, 95% CI: 0.028-0.276, p < 0.001) and anxiety (OR = 0.150, 95% CI: 0.053-0.345, p < 0.001). A higher proportion of smartphone use while alone (vs. with others) was significantly associated with higher odds of depressive symptoms (OR = 3.802, 95% CI: 1.622-11.241, p = 0.006). Time spent on internet browsing was also associated with higher odds of depressive symptoms (OR = 2.663, 95% CI: 1.238-6.843, p = 0.022).

**Table 3.** Multivariate logistic regression models showing associations between mental health symptoms, sociodemographic characteristics, and smartphone use context.

| Independent Variable | Depression |  | Anxiety |  | Suicidal Ideation |  |
| --- | --- | --- | --- | --- | --- | --- |
|  | OR [95% CI] | p-value | OR [95% CI] | p-value | OR [95% CI] | p-value |
| Age (scaled) | 0.721<br>[0.275, 1.805] | 0.490 | 1.438<br>[0.65, 3.249] | 0.370 | 0.654<br>[0.225, 1.566] | 0.383 |
| **Parental education: Post-secondary incomplete/I do not know (ref)** |  |  |  |  |  |  |
| Parental education: Complete post-secondary | 1.42<br>[0.386, 5.395] | 0.598 | 2.598<br>[0.759, 10.101] | 0.143 | 2.702<br>[0.742, 10.91] | 0.142 |
| **Ethnicity: Other (ref)** |  |  |  |  |  |  |
| Ethnicity: European/Canadian | 0.604<br>[0.161, 2.136] | 0.439 | 3.849<br>[1.165, 14.646] | 0.034* | 3.256<br>[0.923, 12.598] | 0.073 |
| **Part-time job: No (ref)** |  |  |  |  |  |  |
| Part-time job status: Yes | 0.529<br>[0.089, 2.988] | 0.469 | 0.083<br>[0.012, 0.425] | 0.005* | 0.039<br>[0.001, 0.367] | 0.015* |
| **Smartphone use context** |  |  |  |  |  |  |
| Proportion of smartphone use surfing the internet | 2.663<br>[1.238, 6.843] | 0.022* | 1.953<br>[0.988, 4.272] | 0.070 | 1.753<br>[0.871, 4.033] | 0.144 |
| Proportion of smartphone solitary vs with others | 3.802<br>[1.622, 11.241] | 0.006* | 1.977<br>[0.997, 4.284] | 0.064 | 1.698<br>[0.817, 3.933] | 0.180 |
| Proportion at home | 0.105<br>[0.028, 0.276] | <0.001* | 0.150<br>[0.053, 0.345] | <0.001* | 0.445<br>[0.191, 0.929] | 0.042<sup>a</sup> |
| **AIC** | 81.859 |  | 87.791 |  | 79.358 |  |

\*Indicates a statistically significant association at the $p < 0.05$ threshold.
<sup>a</sup>Indicates that the association was no longer statistically significant after adjustment for False Discovery Rate

**Table 4** presents adjusted logistic regression models examining associations between smartphone use duration and mental health symptoms. Time spent using smartphones outside the home was significantly associated with depressive symptoms (OR = 4.289, 95% CI: 1.44-16.58, p = 0.018). Part-time employment was also significantly associated with lower odds of suicidal ideation (OR = 0.069, 95% CI: 0.003–0.551, p = 0.030), while no other covariates were statistically significant.

**Table 4.** Multivariate logistic regression models showing associations between mental health symptoms, sociodemographic characteristics, and smartphone use duration.

| Independent variables | Depression |  | Anxiety |  | Suicidal Ideation |  |
| --- | --- | --- | --- | --- | --- | --- |
|  | OR [95% CI] | p-value | OR [95% CI] | p-value | OR [95% CI] | p-value |
| Age (scaled) | 1.295<br>[0.749, 2.283] | 0.356 | 1.437<br>[0.789, 2.711] | 0.243 | 0.849<br>[0.347, 1.842] | 0.695 |
| **Parental education: Post-secondary incomplete/I do not know (ref)** |  |  |  |  |  |  |
| Parental education: Complete post-secondary | 0.610<br>[0.215, 1.647] | 0.336 | 2.891<br>[0.991, 9.141] | 0.059 | 2.065<br>[0.563, 7.883] | 0.275 |
| **Ethnicity: Other (ref)** |  |  |  |  |  |  |
| Ethnicity: European /Canadian | 1.539<br>[0.554, 4.375] | 0.410 | 2.339<br>[0.852, 6.669] | 0.103 | 2.606<br>[0.758, 9.491] | 0.133 |
| **Part-time job: No (ref)** |  |  |  |  |  |  |
| Part-time job status: Yes | 0.879<br>[0.239, 3.116] | 0.842 | 0.424<br>[0.103, 1.592] | 0.215 | 0.069<br>[0.003, 0.551] | 0.030* |

| Smartphone use context |  |  |  |  |  |  |
| --- | --- | --- | --- | --- | --- | --- |
| Time on the internet | 0.308<br>[0.058,<br>1.319] | 0.133 | 1.138<br>[0.258,<br>5.063] | 0.862 | 0.971<br>[0.115,<br>7.799] | 0.977 |
| Time solitary | 1.428<br>[0.632,<br>3.366] | 0.395 | 0.677<br>[0.252,<br>1.645] | 0.403 | 2.066<br>[0.704,<br>7.08] | 0.208 |
| Time outside | 4.289<br>[1.443,<br>16.579] | 0.018* | 2.985<br>[0.998,<br>11.233] | 0.072 | 0.918<br>[0.189,<br>4.721] | 0.915 |
| **AIC** | 116.73 |  | 109.66 |  | 82.167 |  |

\*Indicates a statistically significant association at the $p < 0.05$ threshold.
<sup>a</sup>Indicates that the association was no longer statistically significant after adjustment for the False Discovery Rate

## 4. Discussion

This study investigated how the context of smartphone use, including location, social interaction, activity type, and duration, is associated with mental health symptoms among youth. Framing this within a digital phenotyping approach, we used mEPAs to actively collect high-frequency, in-situ self-reports of behaviour and context through participants’ smartphones [8,10,27]. Although this approach does not involve passive sensing, it can enable the capture of ecologically valid, time-stamped digital data that can function as behavioural markers of mental health [14]. By measuring multiple contextual dimensions rather than screen time alone, this work advances the potential for context-aware digital phenotyping to inform early detection and precision prevention strategies in youth mental health [13,18]. It also builds on the foundation of digital citizen science, where youth are directly engaged in shaping how their data are collected, interpreted, and used – an approach that can improve relevance, transparency, and trust in digital mental health research [9,27].

The results showed that a greater proportion of smartphone use at home was associated with lower odds of anxiety and depressive symptoms, whereas greater duration of smartphone use outside the home was associated with higher odds of depressive symptoms and suicidal ideation. Although no prior studies have, to our knowledge, examined smartphone use in relation to location context, this finding may appear counterintuitive. Previous GPS-based mobility studies often report that spending more time at home is associated with poorer mental health [30,31]. However, GPS mobility metrics quantify total time spent at home regardless of the activities undertaken in that setting [32], whereas our measure captures the proportion of total smartphone use occurring at home, which can differ markedly between individuals with similar overall time at home. Further investigation is warranted to determine whether the association between the proportion of at-home use and mental health varies across levels of total smartphone use. Analyses incorporating an interaction term between proportion and duration, or stratifying models by duration, could clarify whether these associations differ at lower versus higher total use [33]. Additionally, these findings should be interpreted with caution and validated in other populations, as the present analysis was conducted among Canadian youth and may not be generalizable globally.

Beyond physical location, the social context of digital engagement was also associated with mental health outcomes: a greater proportion of smartphone use while alone, compared to use with others, was associated with higher odds of depressive symptoms. This pattern corroborates prior evidence, which found that social use of digital media was negatively associated with poor mental health outcomes in adolescents [34–36]. Digital technologies, including smartphones, can facilitate connection through synchronous and asynchronous communication, which may buffer against psychological distress, particularly when in-person interaction is limited [37]. These findings may seem to contradict studies reporting that social media use is associated with poorer mental health outcomes among adolescents and young adults [38]. However, social media activity is not inherently social; solitary engagement, including behaviours such as passive browsing on platforms designed for interaction, can have very different implications for mental health than active, reciprocal communication. Future work should explore whether the risks associated with solitary digital engagement differ by platform, content type, or frequency, and whether shifting patterns toward more socially connected use can mitigate depressive symptoms. Longitudinal studies and intervention trials are needed to determine the causal pathways linking the social context of smartphone use with mental health outcomes, and to test scalable strategies, such as app features that encourage co-use or group interaction, for integrating protective forms of engagement into daily routines.

Activity type further distinguished associations, as a greater proportion of smartphone time spent on internet surfing, relative to other activities such as texting or gaming, was associated with higher odds of depressive symptoms. This finding aligns with literature showing that habitual or passive online browsing, particularly of algorithm-driven content, is associated with lower well-being and higher depressive symptoms [39–41]. Unlike goal-directed activities (e.g., direct messaging, schoolwork, gaming), internet surfing often involves unstructured engagement with heterogeneous content, which can increase exposure to negative or mood-congruent information [42]. Recent evidence from a community-based sample of adolescents found that exceeding two hours of weekday screen time increased the odds of clinically-relevant anxiety and emotional and behavioural difficulties, with passive scrolling having the strongest negative influence [22]. Future research should examine whether reducing the proportion of smartphone time spent on internet surfing in favour of more interactive or goal-directed activities improves mental health outcomes, and whether interventions at the individual or platform level can shift engagement patterns toward healthier digital behaviours.

Our findings also support the use of digital citizen science as a model for ethical, participatory digital phenotyping. Previous work has demonstrated that co-designing and co-governing digital tools with youth can improve data quality, engagement, and trust [28,43,44]. This is particularly relevant for adolescents, who are both high users of digital technologies and often excluded from shaping how those tools are deployed in health research [16]. Embedding human-centred AI within this model could further ensure that analytic and intervention systems are developed in ways that reflect youth priorities and operate under governance structures they help design [45,46]. Research funders and ethics boards could prioritize youth-led governance structures for digital health projects, ensuring adolescents have formal decision-making roles in how their data are collected and applied [47–49].

Overall, these findings suggest that digital phenotyping strategies incorporating location, social context, and activity type, rather than duration alone, may better identify youth at elevated risk for internalizing symptoms. This approach aligns with emerging models of precision prediction, which use real-time behavioural markers to detect early signs of psychological distress, and precision prevention, which delivers context-specific interventions before symptoms escalate [14,50–52]. Provincial and municipal health systems could embed such multi-context indicators into existing school-based health screening or youth crisis response systems, enabling targeted outreach and support before symptoms progress [53].

## 5. Strengths and Limitations

This study makes an early contribution to the evidence base on context-aware digital phenotyping in youth. Capturing multiple dimensions of smartphone use, including location, social context, activity type, and duration, allowed for a more nuanced understanding of behavioural patterns than total screen time alone, which can inform more targeted and effective interventions. Digital deployment demonstrated feasibility outside health systems, an important step toward integrating such methods into routine youth mental health monitoring at scale [54]. Another strength was the integration of citizen science principles [15,27,29], making this, to our knowledge, the first study to combine mEPAs with active youth involvement to examine multiple mental health outcomes. Engaging youth citizen scientists enabled measures that were clearer, relevant, and acceptable, which likely improved engagement and data quality. Nevertheless, future research should prioritize the development of validated tools to systematically evaluate the citizen science approach in digital phenotyping studies, as no standard metrics currently exist to assess its impact on data validity.

Despite these strengths, the present study had several limitations. The study’s generalizability is limited, as participants were drawn from a small number of schools in one Canadian city; future research should recruit more diverse samples across multiple cultural and geographic contexts. Another limitation is that certain covariates, such as income and gender identity, were not included in the analyses because of small cell counts, which could increase residual confounding [55]; future studies with larger sample sizes are needed to enable inclusion of these variables. Additionally, while the Generalized Anxiety Disorder-2 instrument is validated in youth and single-item depression and suicidal ideation measures have precedent in comparable studies, these instruments have not been formally validated in digital phenotyping contexts [56]. These design choices prioritized feasibility for early-stage evidence generation but highlight the need for replication in larger, more diverse samples with robust digital mental health measures before informing policy or clinical practice.

## 6. Conclusion

This study demonstrates that context-aware digital phenotyping, particularly smartphone use patterns such as location, social setting, and activity type, provides meaningful insights into youth mental health. By leveraging mEPAs, it advances the utility of real-time, youth-informed digital surveillance for early identification of mental health symptoms. While the sample size and generalizability are limited, the findings highlight the shortcomings of smartphone use as a standalone metric and support a shift toward more nuanced, context-sensitive approaches. This work lays a foundation for scalable, participatory, and ethical models of digital mental health monitoring, particularly in underserved settings, aligned with emerging frameworks of precision prediction and prevention.

## Data Availability

All data produced in the present study are available upon reasonable request to the authors

## References

[1] Kieling C, Buchweitz C, Caye A, Silvani J, Ameis SH, Brunoni AR, et al. Worldwide Prevalence and Disability From Mental Disorders Across Childhood and Adolescence: Evidence From the Global Burden of Disease Study. JAMA Psychiatry 2024;81:347–56. 10.1001/jamapsychiatry.2023.5051.

[2] GBD 2019 Mental Disorders Collaborators. Global, regional, and national burden of 12 mental disorders in 204 countries and territories, 1990–2019: a systematic analysis for the Global Burden of Disease Study 2019. The Lancet Psychiatry 2022;9:137–50. 10.1016/S2215-0366(21)00395-3.

[3] Ghafari M, Nadi T, Bahadivand-Chegini S, Doosti-Irani A. Global prevalence of unmet need for mental health care among adolescents: A systematic review and meta-analysis. Archives of Psychiatric Nursing 2022;36:1–6. 10.1016/j.apnu.2021.10.008.

[4] Arias D, Saxena S, Verguet S. Quantifying the global burden of mental disorders and their economic value. eClinicalMedicine 2022;54. 10.1016/j.eclinm.2022.101675.

[5] McGorry PD, Mei C, Dalal N, Alvarez-Jimenez M, Blakemore S-J, Browne V, et al. The Lancet Psychiatry Commission on youth mental health. The Lancet Psychiatry 2024;11:731–74. 10.1016/S2215-0366(24)00163-9.

[6] Teckchandani TA, Shields RE, Andrews KL, Nisbet J, Afifi TO, Asmundson GJG, et al. Monthly Mental Health Self-Monitoring and Positive Changes in Mental Health Disorder Symptoms Scores Among Royal Canadian Mounted Police Cadets. Int J Cogn Behav Ther 2025. 10.1007/s41811-025-00231-w.

[7] Barch DM, Albaugh MD, Baskin-Sommers A, Bryant BE, Clark DB, Dick AS, et al. Demographic and mental health assessments in the adolescent brain and cognitive development study: Updates and age-related trajectories. Developmental Cognitive Neuroscience 2021;52:101031. 10.1016/j.dcn.2021.101031.

[8] Ibrahim ST, Hammami N, Katapally TR. Traditional surveys versus ecological momentary assessments: Digital citizen science approaches to improve ethical physical activity surveillance among youth. PLOS Digit Health 2023;2:e0000294. 10.1371/journal.pdig.0000294.

[9] Ibrahim ST, Patel J, Katapally TR. Digital citizen science for ethical monitoring of youth physical activity frequency: Comparing mobile ecological prospective assessments and retrospective recall. PLOS Digit Health 2025;4:e0000840. 10.1371/journal.pdig.0000840.

[10] Al-Akshar S, Tolulope Ibrahim S, Katapally TR. How can digital citizen science approaches improve ethical smartphone use surveillance among youth: Traditional surveys versus ecological momentary assessments. PLOS Digit Health 2024;3:e0000448. 10.1371/journal.pdig.0000448.

[11] Li M, Patel J, Katapally TR. Towards ethical surveillance of smartphone use among youth: exploratory digital citizen science approaches shaping the understanding of ubiquitous technology use. Technology in Society 2025;83:103012. 10.1016/j.techsoc.2025.103012.

[12] Torous J, Bucci S, Bell IH, Kessing LV, Faurholt-Jepsen M, Whelan P, et al. The growing field of digital psychiatry: current evidence and the future of apps, social media, chatbots, and virtual reality. World Psychiatry 2021;20:318–35. 10.1002/wps.20883.

[13] Bufano P, Laurino M, Said S, Tognetti A, Menicucci D. Digital Phenotyping for Monitoring Mental Disorders: Systematic Review. Journal of Medical Internet Research 2023;25:e46778. 10.2196/46778.

[14] Patel J, Hung C, Katapally TR. Evaluating predictive artificial intelligence approaches used in mobile health platforms to forecast mental health symptoms among youth: a systematic review. Psychiatry Research 2025;343:116277. 10.1016/j.psychres.2024.116277.

[15] Katapally TR. A Global Digital Citizen Science Policy to Tackle Pandemics Like COVID-19. J Med Internet Res 2020;22:e19357. 10.2196/19357.

[16] Wike R, Silver L, Fetterolf J, Huang C, Austin S, Clancy L, et al. Social media seen as mostly good for democracy across many nations, but US is a major outlier. vol. 6. Pew Research Center Washington, DC, USA; 2022.

[17] Beames JR, Han J, Shvetcov A, Zheng WY, Slade A, Dabash O, et al. Use of smartphone sensor data in detecting and predicting depression and anxiety in young people (12–25 years): A scoping review. Heliyon 2024;10:e35472. 10.1016/j.heliyon.2024.e35472.

[18] Currey D, Torous J. Digital Phenotyping Data to Predict Symptom Improvement and Mental Health App Personalization in College Students: Prospective Validation of a Predictive Model. Journal of Medical Internet Research 2023;25:e39258. 10.2196/39258.

[19] Langholm C, Alon N, Perret S, Torous J. Risk scores in digital psychiatry: Expanding the reach of complex smartphone data by condensing it into simple results. Journal of Behavioral and Cognitive Therapy 2023;33:90–6. 10.1016/j.jbct.2023.05.004.

[20] Sano A, Taylor S, McHill AW, Phillips AJ, Barger LK, Klerman E, et al. Identifying Objective Physiological Markers and Modifiable Behaviors for Self-Reported Stress and Mental Health Status Using Wearable Sensors and Mobile Phones: Observational Study. J Med Internet Res 2018;20:e210. 10.2196/jmir.9410.

[21] Jafarlou S, Lai J, Azimi I, Mousavi Z, Labbaf S, Jain RC, et al. Objective Prediction of Next-Day’s Affect Using Multimodal Physiological and Behavioral Data: Algorithm Development and Validation Study. JMIR Formative Research 2023;7:e39425. 10.2196/39425.

[22] Choi EJ, Christiaans E, Duerden EG. Screen time woes: Social media posting, scrolling, externalizing behaviors, and anxiety in adolescents. Computers in Human Behavior 2025;170:108688. 10.1016/j.chb.2025.108688.

[23] Latif MZ, Hussain I, Saeed R, Qureshi MA, Maqsood U. Use of Smart Phones and Social Media in Medical Education: Trends, Advantages, Challenges and Barriers. Acta Inform Med 2019;27:133–8. 10.5455/aim.2019.27.133-138.

[24] Brogly C, Lizotte DJ, Mitchell M, Speechley M, MacDougall A, Huner E, et al. An app-based ecological momentary assessment of undergraduate student mental Health during the COVID-19 pandemic in Canada (Smart Healthy Campus Version 2.0): Longitudinal study. PLOS Digital Health 2024;3:e0000239. 10.1371/journal.pdig.0000239.

[25] Portillo-Van Diest A, Mortier P, Ballester L, Amigo F, Carrasco P, Falcó R, et al. Ecological Momentary Assessment of Mental Health Problems Among University Students: Data Quality Evaluation Study. J Med Internet Res 2024;26:e55712. 10.2196/55712.

[26] Hammoud R, Tognin S, Smythe M, Gibbons J, Davidson N, Bakolis I, et al. Smartphone-based ecological momentary assessment reveals an incremental association between natural diversity and mental wellbeing. Sci Rep 2024;14:7051. 10.1038/s41598-024-55940-7.

[27] Katapally TR, Chu LM. Digital epidemiological and citizen science methodology to capture prospective physical activity in free-living conditions: a SMART Platform study. BMJ Open 2020;10:e036787. 10.1136/bmjopen-2020-036787.

[28] Katapally TR, Hammami N, Chu LM. A randomized community trial to advance digital epidemiological and mHealth citizen scientist compliance: A smart platform study. PLOS ONE 2021;16:e0259486. 10.1371/journal.pone.0259486.

[29] Katapally TR, Bhawra J, Leatherdale ST, Ferguson L, Longo J, Rainham D, et al. The SMART Study, a Mobile Health and Citizen Science Methodological Platform for Active Living Surveillance, Integrated Knowledge Translation, and Policy Interventions: Longitudinal Study. JMIR Public Health and Surveillance 2018;4:e8953. 10.2196/publichealth.8953.

[30] Laiou P, Kaliukhovich DA, Folarin AA, Ranjan Y, Rashid Z, Conde P, et al. The Association Between Home Stay and Symptom Severity in Major Depressive Disorder: Preliminary Findings From a Multicenter Observational Study Using Geolocation Data From Smartphones. JMIR Mhealth Uhealth 2022;10:e28095. 10.2196/28095.

[31] Depp CA, Bashem J, Moore RC, Holden JL, Mikhael T, Swendsen J, et al. GPS mobility as a digital biomarker of negative symptoms in schizophrenia: a case control study. NPJ Digit Med 2019;2:108. 10.1038/s41746-019-0182-1.

[32] Katapally TR, Bhawra J, Patel P. A systematic review of the evolution of GPS use in active living research: A state of the evidence for research, policy, and practice. Health & Place 2020;66:102453. 10.1016/j.healthplace.2020.102453.

[33] Greenland S. Effect Modification and Interaction. Wiley StatsRef: Statistics Reference Online, John Wiley & Sons, Ltd; 2015, p. 1–5. 10.1002/9781118445112.stat03728.pub2.

[34] Rudolf R, Kim N. Smartphone use, gender, and adolescent mental health: Longitudinal evidence from South Korea. SSM - Population Health 2024;28:101722. 10.1016/j.ssmph.2024.101722.

[35] Frison E, Eggermont S. Exploring the Relationships Between Different Types of Facebook Use, Perceived Online Social Support, and Adolescents’ Depressed Mood. Social Science Computer Review 2016;34:153–71. 10.1177/0894439314567449.

[36] Abi-Jaoude E, Naylor KT, Pignatiello A. Smartphones, social media use and youth mental health. CMAJ 2020;192:E136–41. 10.1503/cmaj.190434.

[37] Juvonen J, Schacter HL, Lessard LM. Connecting electronically with friends to cope with isolation during COVID-19 pandemic. Journal of Social and Personal Relationships 2021;38:1782–99. 10.1177/0265407521998459.

[38] Khalaf AM, Alubied AA, Khalaf AM, Rifaey AA. The Impact of Social Media on the Mental Health of Adolescents and Young Adults: A Systematic Review. Cureus n.d.;15:e42990. 10.7759/cureus.42990.

[39] Taskin S, Yildirim Kurtulus H, Satici SA, Deniz ME. Doomscrolling and mental well-being in social media users: A serial mediation through mindfulness and secondary traumatic stress. J Community Psychol 2024;52:512–24. 10.1002/jcop.23111.

[40] Shabahang R, Hwang H, Thomas EF, Aruguete MS, McCutcheon LE, Orosz G, et al. Doomscrolling evokes existential anxiety and fosters pessimism about human nature? Evidence from Iran and the United States. Computers in Human Behavior Reports 2024;15:100438. 10.1016/j.chbr.2024.100438.

[41] Satici SA, Gocet Tekin E, Deniz ME, Satici B. Doomscrolling Scale: its Association with Personality Traits, Psychological Distress, Social Media Use, and Wellbeing. Appl Res Qual Life 2023;18:833–47. 10.1007/s11482-022-10110-7.

[42] Kelly CA, Sharot T. Web-browsing patterns reflect and shape mood and mental health. Nat Hum Behav 2025;9:133–46. 10.1038/s41562-024-02065-6.

[43] Katapally TR, Elsahli N, Bhawra J. DiScO: novel rapid systems mapping to inform digital transformation of health systems. Front Public Health 2024;12. 10.3389/fpubh.2024.1441328.

[44] Katapally TR, Elsahli N, Ibrahim ST, Bhawra J. Human-Centered Digital Nudging to Promote Youth Mental Health: A Serendipitous Natural Experiment Enabled by a Digital Health Platform 2024. 10.2139/ssrn.4819875.

[45] Schmager S, Pappas IO, Vassilakopoulou P. Understanding Human-Centred AI: a review of its defining elements and a research agenda. Behaviour & Information Technology n.d.;0:1–40. 10.1080/0144929X.2024.2448719.

[46] Chen Y, Clayton EW, Novak LL, Anders S, Malin B. Human-Centered Design to Address Biases in Artificial Intelligence. J Med Internet Res 2023;25:e43251. 10.2196/43251.

[47] Piasecki J, Cheah PY. Ownership of individual-level health data, data sharing, and data governance. BMC Medical Ethics 2022;23:104. 10.1186/s12910-022-00848-y.

[48] Bowie R. Indigenous Self-Governance and the Deployment of Knowledge in Collaborative Environmental Management in Canada. Journal of Canadian Studies 2013;47:91–121. 10.3138/jcs.47.1.91.

[49] Abbas AE, van Velzen T, Ofe H, van de Kaa G, Zuiderwijk A, de Reuver M. Beyond control over data: Conceptualizing data sovereignty from a social contract perspective. Electron Markets 2024;34:20. 10.1007/s12525-024-00695-2.

[50] Ibrahim ST, Li M, Patel J, Katapally TR. Utilizing natural language processing for precision prevention of mental health disorders among youth: A systematic review. Computers in Biology and Medicine 2025;188:109859. 10.1016/j.compbiomed.2025.109859.

[51] Pearson TA, Vitalis D, Pratt C, Campo R, Armoundas AA, Au D, et al. The Science of Precision Prevention. JACC Adv 2023;3:100759. 10.1016/j.jacadv.2023.100759.

[52] Fernandes BS, Williams LM, Steiner J, Leboyer M, Carvalho AF, Berk M. The new field of ‘precision psychiatry.’ BMC Medicine 2017;15:80. 10.1186/s12916-017-0849-x.

[53] Katapally TR, Ibrahim ST. Digital Health Dashboards for Decision-Making to Enable Rapid Responses During Public Health Crises: Replicable and Scalable Methodology. JMIR Research Protocols 2023;12:e46810. 10.2196/46810.

[54] Patel V, Saxena S, Lund C, Thornicroft G, Baingana F, Bolton P, et al. The Lancet Commission on global mental health and sustainable development. Lancet 2018;392:1553–98. 10.1016/S0140-6736(18)31612-X.

[55] Bhopal RS. Error, bias, and confounding in epidemiology. In: Bhopal RS, editor. Concepts of Epidemiology: Integrating the ideas, theories, principles, and methods of epidemiology, Oxford University Press; 2016, p. 0. 10.1093/med/9780198739685.003.0004.

[56] Sapra A, Bhandari P, Sharma S, Chanpura T, Lopp L. Using Generalized Anxiety Disorder-2 (GAD-2) and GAD-7 in a Primary Care Setting. Cureus n.d.;12:e8224. 10.7759/cureus.8224.

